# Peripheral blood profiles reflecting progenitor lineage balance predict treatment response in chronic myeloid leukemia

**DOI:** 10.64898/2026.01.15.26344146

**Authors:** Kohjin Suzuki, Naoki Watanabe, Yutaka Tsukune, Tadaaki Inano, Shintaro Kinoshita, Kazuhiro Yamada, Miki Ando, Tomoiku Takaku

## Abstract

Early achievement of deep remission improves patients’ outcome in chronic myeloid leukemia (CML) treatment, highlighting the need for predictive indicators before therapy initiation. This study aimed to develop a tool to predict CML treatment responses to guide optimal therapy selection. Using hierarchical clustering of complete blood count (CBC) data at diagnosis, patients were stratified into two clusters. Patients in Cluster 1 had higher *BCR::ABL1*^IS^ mRNA levels at 3 and 6 months post-treatment and lower rates of major molecular response compared to cluster 2. Cluster 1 also showed increased granulocyte and immature white blood cell counts and decreased erythroid parameters. Flow cytometric analysis of bone marrow mononuclear cells revealed that cluster 1 had a significant increase in hematopoietic stem cell fractions and a higher ratio of granulocyte-macrophage progenitors to megakaryocyte-erythroid progenitors compared to cluster 2. These findings suggest that differences in bone marrow progenitor cell differentiation affect peripheral blood profiles. Artificial intelligence-driven ghost cytometry (GC) was evaluated for its ability to comprehensively capture these changes and successfully distinguished patients with poorer treatment responses, with GC scores at diagnosis strongly correlating with *BCR::ABL1*^IS^ mRNA levels at 3 and 6 months post-treatment initiation. The study indicates that multivariate analysis of CBC or GC analysis may enable simple, early prediction of CML treatment efficacy, potentially contributing to effective and individualized CML therapy.

## Introduction

Chronic myeloid leukemia (CML) is a myeloproliferative neoplasm characterized by a marked proliferation of granulocytic lineage cells, resulting from the formation of the Philadelphia chromosome due to a translocation between chromosomes 9 and 22 in hematopoietic stem cells (HSCs)^1,2^. This leads to the expression of the *BCR::ABL1* fusion gene^3^. The treatment of CML involves the use of tyrosine kinase inhibitors (TKIs) targeting *BCR::ABL1*^4^, and the introduction of TKIs has dramatically improved the prognosis of patients with CML^5^. The therapeutic milestone of TKI treatment include achieving a partial cytogenetic response (PCyR) within 3 months, a complete cytogenetic response (CCyR) within 6 months, and a major molecular response (MMR) within 12 months, with the ultimate aim of attaining a deep molecular response (DMR)^6,7^. However, various adverse events (AEs) associated with long-term TKI administration have been reported including fatal cardiovascular AEs^8^, which can negatively impact the quality of life of CML patients^9^. Currently, the European LeukemiaNet (ELN) 2025 recommendations advocate for the attempt of treatment-free remission (TFR) in patients who have achieved sufficient therapeutic response, making TFR as one of the treatment goals^7^. Nevertheless, the success rate of TFR is approximately 50% in clinical trials^10,11^ and about half of that in real-world clinical practice. Recent studies have shown that patients who achieve deep molecular response early in the course of treatment have a higher rate of successful TFR^12^, highlighting the importance of predicting individual treatment responsiveness and initiating therapy with the appropriate type and dose of TKI. Although prognostic scoring systems such as the EUTOS long-term survival (ELTS) score exist^13^, tools for predicting treatment response at diagnosis remain under investigation.

One of the main causes of TFR failure is the persistence of leukemic stem cells (LSCs) after treatment^14^. In other words, the status of hematopoietic stem and progenitor cells (HSPCs) in CML patients is considered to have a significant impact on the achievement of TFR. HSCs differentiate into multipotent progenitors (MPPs), which then branch into common lymphoid progenitors (CLPs) and common myeloid progenitors (CMPs). CMPs further differentiate into granulocyte-macrophage progenitors (GMPs) and megakaryocyte-erythroid progenitors (MEPs). CLPs differentiate into various lymphocytes and NK cells, GMPs differentiate into myeloid lineage cells such as granulocytes, and MEPs differentiate into erythroblasts and megakaryocytes^15,16^. Abnormal differentiation of these HSPCs is considered to be associated with treatment resistance and disease progression^17^, making them important factors in the pathophysiological understanding of CML. However, bone marrow aspiration can cause substantial discomfort and inconvenience to patients, and frequent flow cytometric or genetic analyses of bone marrow cells are not feasible. Therefore, there is a need for a simple and convenient tool to assess the status of bone marrow HSPCs.

In this study, we aimed to develop a predictive tool for CML treatment responses to support appropriate therapeutic selection to improve the achievement rate of TFR.

## Methods

### Clinical samples

Peripheral blood and bone marrow samples were obtained from 12 patients with CML at diagnosis. Patients’ characteristics were shown in Table 1. During the 6-month observation period, all patients received initial treatment with second-generation TKIs, and subsequent therapy was selected from alternative second-, third-generation TKIs, or asciminib. This study was approved by the Ethics Committee of Juntendo University School of Medicine and Sysmex Corporation and adhered to the Declaration of Helsinki. Patients provided informed consent by accessing the study information published on the website and were free to withdraw their consent at any point during the study, as per the Ethical Guidelines for Medical and Health Research Involving Human Subjects. The ethics committee waived the requirement for written informed consent. Clinical information and data pertaining to clinical tests, including the international scale (IS) of *BCR::ABL1*^IS^ mRNA, were retrieved from medical records.

**Table 1.**
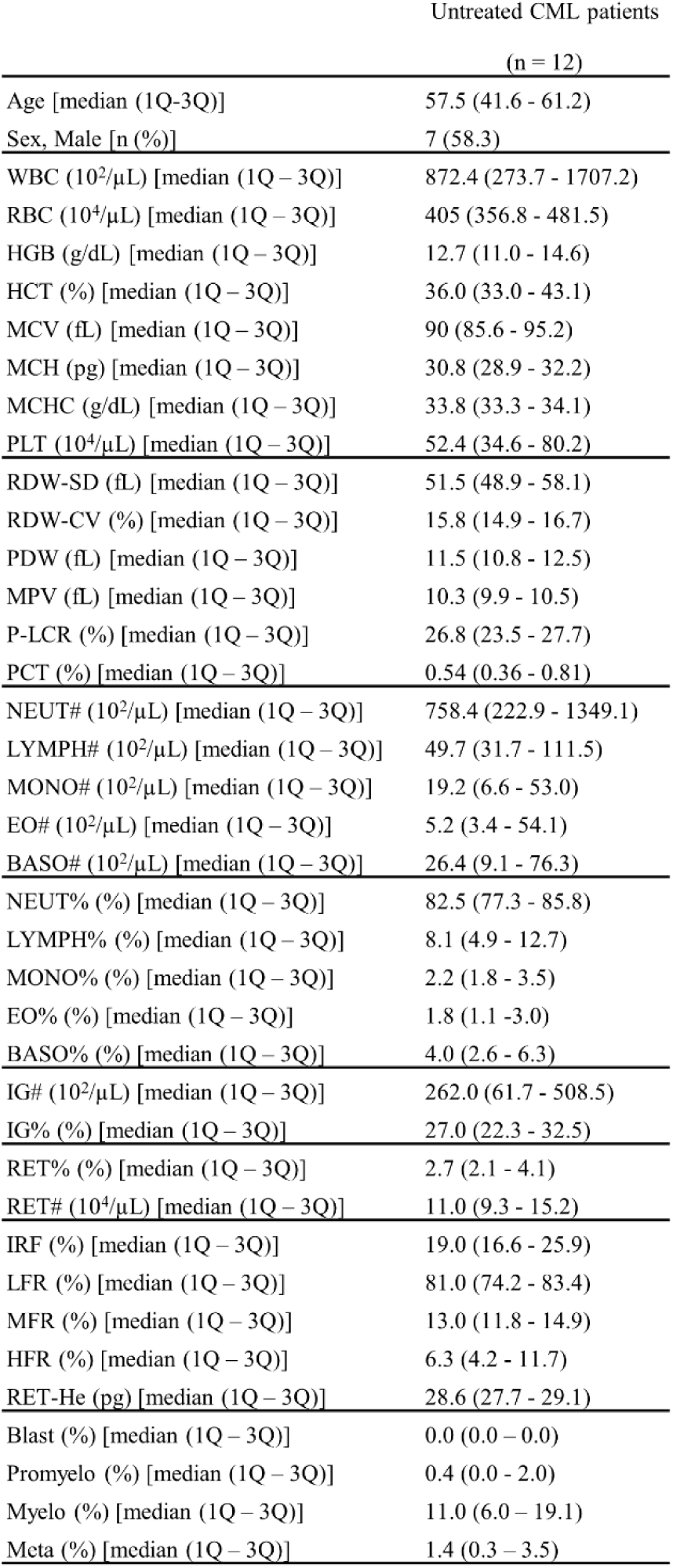
Patients’ characteristics and baseline hematological parameters of the study cohort at diagnosis.

### Complete blood count

Complete blood count data were obtained using an automated hematology analyzer XN-450 (Sysmex, Hyogo, Japan). The following information was obtained: WBC, white blood cell; RBC, red blood cell; HGB, hemoglobin; HCT, hematocrit; MCV, mean corpuscular volume; MCH, mean corpuscular hemoglobin; MCHC, mean corpuscular hemoglobin concentration; PLT, platelet; RDW-SD, standard deviation of red cell distribution width; RDW-CV, coefficient of variation of red cell distribution width; PDW, platelet distribution width; P-LCR, platelet large cell ratio; PCT, plateletcrit; NEUT, neutrophil; LYMPH, lymphocyte; MONO, monocytes; EO, eosinophil; BASO, basophil; IG, immature granulocyte; RET, reticulocyte; IRF, immature reticulocyte fraction; LFR, low-fluorescence reticulocyte; MFR, middle-fluorescence reticulocyte; HFR, high-fluorescence reticulocyte; and RET-He, reticulocyte hemoglobin equivalent. Blast, promyelocyte, myelocyte, and metamyelocyte were obtained from the medical record.

### Flow cytometry

Mononuclear cells in bone marrow aspirates obtained from CML patients were extracted using LymphoSep (MP Biomedicals, Solon, OH, USA), followed by cryopreservation using CellBanker1 (Nippon Zenyaku, Fukushima, Japan) until use. Cryopreserved bone marrow mononuclear cells (BM-MNCs) were thawed in 37℃ water bath for 5 minutes, followed by phosphate-buffered saline wash. Thawed BM-MNCs were filtered to remove cell aggregates and subsequently stained with fluorochrome-conjugated antibodies for 30 minutes on ice for flow cytometric analysis using FACSCelesta (Becton Dickinson, Franklin Lakes, NJ, USA). The data related to the flow cytometry measurements obtained in this study were analyzed using the FlowJo software (version 10.8.1, Becton Dickinson). The antibodies used were anti-CD38 V450 (BD Biosciences, Franklin Lakes, NJ, USA, Cat# 646851), anti-CD45 V500-C (BD Biosciences, Cat# 647449), anti-CD10 FITC (BD Biosciences, Cat# 340925), anti-CD135 PE (BD Biosciences, Cat# 558996), anti-CD34 PE-Cy7 (BD Biosciences, Cat# 348791), anti-CD90 APC (BD Biosciences, Cat# 559869), and anti-CD45RA APC-H7 (BD Biosciences, Cat# 560674). Viable and dead cells were distinguished using 7-aminoactinomycin D (7-AAD, BD Biosciences, Cat# 559925) staining.

### Artificial intelligence-driven flow cytometry analysis using ghost cytometry

The ghost cytometry (GC) measurements analyzed in this study were derived from data previously acquired in our earlier studies^18^, which was performed using a prototype device developed by Sysmex Corporation (Hyogo, Japan). The measurement and analysis procedures were conducted according to the protocols described in our previous publication^18^. The discrimination performance between two cell types using GC was evaluated using the F1 score, which serves as an indicator of analytical accuracy. The F1 score was calculated as the harmonic means of precision and recall.

Precision was defined as the ratio of true positives to the sum of true positives and false positives, whereas recall was defined as the ratio of true positives to the sum of true positives and false negatives. The F1 score ranged from 0 to 1 (0 to 100%), with higher values indicating greater discriminative ability between the two cell groups.

### Statistical analysis

For statistical analyses, the Mann–Whitney U test was used to compare differences between two experimental groups, and the Fisher’s exact test was used to compare differences in proportions between the two experimental groups. All statistical tests were two-tailed, and statistical significance was set at p < 0.05. Spearman’s rank correlation coefficient was used to evaluate the correlation between two factors. Unsupervised hierarchical cluster analysis was performed using Cluster 3.0 (University of Tokyo Human Genome Center). Cluster analysis was performed using complete linkage based on Euclidean distance. Statistical analyses were performed using EZR, which is based on R and R commander^19^.

## Results

### Complete blood count comparison between patient groups with different treatment responses

To identify factors reflecting the therapeutic efficacy in CML patients, we first analyzed 12 cases of CML. *BCR::ABL1*^IS^ mRNA levels were obtained from medical records over time from diagnosis to 6 months after TKI initiation. Patients with *BCR::ABL1*^IS^ mRNA ≤1.5% at 3 months after treatment initiation were defined as early responders (n = 7), while those with *BCR::ABL1*^IS^ mRNA >1.5% were defined as late responders (n = 5) (Figure 1A). At both 3 and 6 months after treatment, *BCR::ABL1*^IS^ mRNA levels were significantly higher in the late responders compared to the early responders (Figure 1B, p < 0.01 and < 0.05, respectively). In terms of treatment response, 60% of late responders were classified as failure at 3 months and 40% at 6 months, whereas all early responders achieved at least CCyR at 3 months and at least MMR at 6 months (Figure 1C).

**Figure 1.**
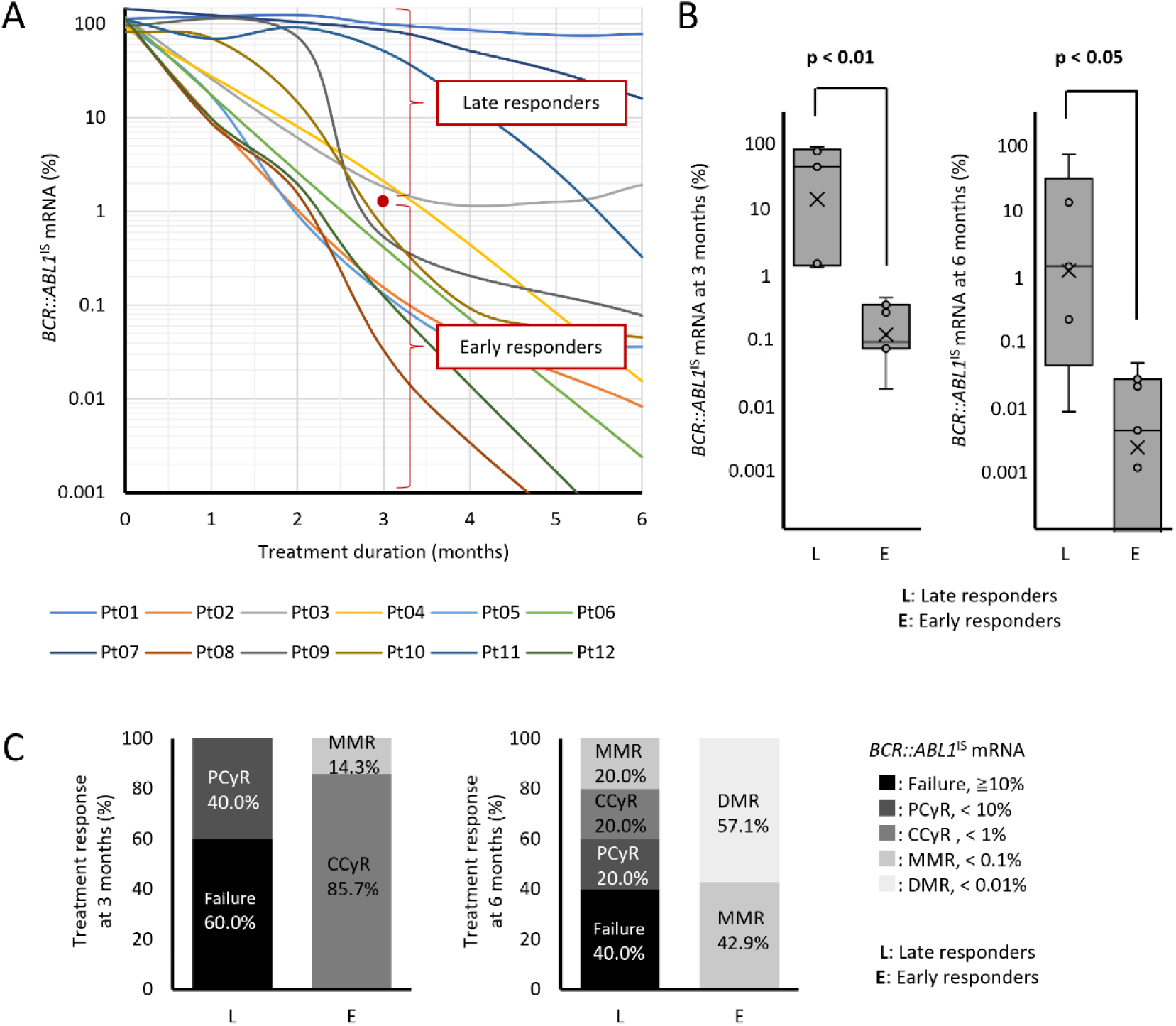
Definition of late and early responders in this study. A. Changes in *BCR::ABL1*^IS^ mRNA during TKI treatment. Patients with *BCR::ABL1*^IS^ mRNA levels of 1.5% or higher at three months of treatment initiation were defined as late responder (n=5), while those with levels below 1.5% were defined as early responder (n=7). B. Comparison of *BCR::ABL1*^IS^ mRNA levels between the two groups at three months and six months after the treatment initiation. C. Treatment responses between late and early responders. PCyR, Partial Cytogenetic Response; CCyR, Complete Cytogenetic Response; MMR, Major Molecular Response; DMR, Deep Molecular Response; Failure, those who did not achieve PCyR.

Next, a comparison of the complete blood count (CBC) profiles at diagnosis between the two groups revealed that the late responders had significantly higher white blood cell (WBC) count, neutrophil count, monocyte count, basophil count, and immature granulocyte (IG) count than the early responders (Table 2). IGs are defined as early precursors of granulocytic white blood cells, including promyelocytes, myelocytes, and metamyelocytes^20^. These findings suggest that the CBC profile at diagnosis differs between groups with distinct treatment responses.

**Table 2.** Comparison of CBC data at diagnosis between late and early responders.

|  | Late responders<br>(n = 5) | Early responders<br>(n = 7) | P value |
| --- | --- | --- | --- |
| Age [median (1Q-3Q)] | 61.0 (42.5 – 64.0) | 56.0 (41.0 – 62.0) | 0.83 |
| Sex, Male [n (%)] | 1 (20.0) | 5 (71.4) | NA |
| WBC ($10^2/\mu\text{L}$) [median (1Q – 3Q)] | 1730.5 (952.6 – 2629.7) | 417.6 (197.1 – 1014.3) | 0.02 |
| RBC ($10^4/\mu\text{L}$) [median (1Q – 3Q)] | 377.0 (317.0 – 412.5) | 424.0 (383.0 – 507.0) | 0.17 |
| HGB (g/dL) [median (1Q – 3Q)] | 11.4 (10.1 – 13.7) | 13.1 (11.6 – 14.8) | 0.35 |
| HCT (%) [median (1Q – 3Q)] | 33.6 (29.9 – 38.8) | 39.4 (34.2 – 44.1) | 0.24 |
| MCV (fL) [median (1Q – 3Q)] | 93.7 (87.8 – 98.2) | 88.9 (85.1 – 95.2) | 0.31 |
| MCH (pg) [median (1Q – 3Q)] | 32.4 (30.0 – 33.9) | 30.8 (28.9 – 32.2) | 0.07 |
| MCHC (g/dL) [median (1Q – 3Q)] | 34.1 (33.6 – 35.3) | 33.5 (33.2 – 33.9) | 0.12 |
| PLT ($10^4/\mu\text{L}$) [median (1Q – 3Q)] | 56.3 (39.7 – 72.4) | 46.7 (34.2 – 81.0) | 0.74 |
| RDW-SD (fL) [median (1Q – 3Q)] | 54.6 (50.3 – 57.6) | 49.4 (46.0 – 58.7) | 0.89 |
| RDW-CV (%) [median (1Q – 3Q)] | 16.0 (15.2 – 16.6) | 15.1 (14.8 – 16.9) | 0.92 |
| PDW (fL) [median (1Q – 3Q)] | 11.2 (10.0 – 12.6) | 11.8 (10.9 – 12.5) | 0.58 |
| MPV (fL) [median (1Q – 3Q)] | 10.1 (9.6 – 10.7) | 10.3 (10.0 – 10.5) | 0.83 |
| P-LCR (%) [median (1Q – 3Q)] | 25.3 (21.1 – 29.9) | 26.9 (25.4 – 27.8) | 0.74 |
| PCT (%) [median (1Q – 3Q)] | 0.6 (0.4 – 0.8) | 0.5 (0.4 – 0.8) | 0.79 |
| NEUT# ($10^2/\mu\text{L}$) [median (1Q – 3Q)] | 1324.2 (778.2 – 2353.6) | 354.1 (142.4 – 854.5) | 0.03 |
| LYMPH# ($10^2/\mu\text{L}$) [median (1Q – 3Q)] | 103.1 (47.9 – 129.2) | 33.6 (30.8 – 77.3) | 0.09 |
| MONO# ($10^2/\mu\text{L}$) [median (1Q – 3Q)] | 47.4 (30.3 – 91.1) | 11.8 (3.7 – 15.1) | 0.02 |
| EO# ($10^2/\mu\text{L}$) [median (1Q – 3Q)] | 41.0 (18.0 – 58.8) | 4.0 (2.9 – 4.6) | 0.11 |
| BASO# ($10^2/\mu\text{L}$) [median (1Q – 3Q)] | 76.9 (43.8 – 100.5) | 10.3 (8.0 – 30.8) | 0.01 |
| NEUT% (%) [median (1Q – 3Q)] | 81.9 (78.9 – 89.1) | 83.0 (72.2 – 84.8) | 0.45 |
| LYMPH% (%) [median (1Q – 3Q)] | 6.9 (3.3 – 8.3) | 11.3 (5.5 – 13.7) | 0.11 |
| MONO% (%) [median (1Q – 3Q)] | 2.8 (1.7 – 7.0) | 2.1 (1.9 – 3.3) | 0.15 |
| EO% (%) [median (1Q – 3Q)] | 2.6 (1.8 – 4.4) | 1.2 (0.6 – 3.0) | 0.20 |
| BASO% (%) [median (1Q – 3Q)] | 4.9 (3.2 – 13.6) | 3.5 (2.5 – 4.5) | 0.30 |
| IG# ($10^2/\mu\text{L}$) [median (1Q – 3Q)] | 437.3 (257.5 – 978.3) | 99.4 (9.0 – 287.8) | 0.04 |
| IG% (%) [median (1Q – 3Q)] | 27.2 (26.0 – 37.4) | 23.8 (6.7 – 32.3) | 0.13 |
| RET% (%) [median (1Q – 3Q)] | 2.7 (1.9 – 4.4) | 2.7 (2.0 – 4.3) | 0.95 |
| RET# ($10^4/\mu\text{L}$) [median (1Q – 3Q)] | 10.0 (6.4 – 16.2) | 12.0 (9.9 – 15.5) | 0.55 |
| IRF (%) [median (1Q – 3Q)] | 23.4 (18.7 – 30.0) | 18.9 (13.5 – 24.8) | 0.51 |
| LFR (%) [median (1Q – 3Q)] | 76.6 (70.0 – 81.4) | 81.1 (75.2 – 86.5) | 0.51 |
| MFR (%) [median (1Q – 3Q)] | 14.4 (12.6 – 15.2) | 12.6 (10.0 – 14.1) | 0.19 |
| HFR (%) [median (1Q – 3Q)] | 11.5 (4.5 – 15.2) | 6.1 (1.8 – 10.7) | 0.68 |
| RET-He (pg) [median (1Q – 3Q)] | 28.4 (27.4 – 29.9) | 28.8 (27.7 – 29.2) | 0.97 |
| Blast (%) [median (1Q – 3Q)] | 0.0 (0.0 – 0.0) | 0.0 (0.0 – 0.0) | NA |
| Promyelo (%) [median (1Q – 3Q)] | 1.2 (0.4 – 3.7) | 0.0 (0.0 – 0.6) | 0.11 |
| Myelo (%) [median (1Q – 3Q)] | 15.8 (8.9 – 33.7) | 7.0 (6.0 – 18.0) | 0.12 |
| Meta (%) [median (1Q – 3Q)] | 2.0 (0.4 – 4.0) | 1.4 (0.0 – 3.2) | 1.00 |

### Clustering analysis using complete blood count data classified patients with different treatment responses

Since the various CBC parameters at diagnosis appeared to be effective for classifying CML patients according to subsequent treatment response, we next performed hierarchical clustering analysis using those CBC data, stratifying patients into cluster 1 (n = 8) and cluster 2 (n = 4) (Figure 2A). There were no significant differences in age, sex, or type of first-line TKI between the two clusters (Figure 2B). However, *BCR::ABL1*^IS^ mRNA levels at 3 and 6 months after treatment initiation were significantly higher in cluster 1 compared to cluster 2 (Figure 2C, p < 0.05 for both). In terms of treatment response, 60% of patients in cluster 1 were classified as failure at 3 months and 25% at 6 months, whereas all patients in cluster 2 achieved at least CCyR at 3 months and at least MMR at 6 months (Figure 2D). Comparison of each CBC parameters revealed that various white blood cell counts were significantly higher in cluster 1, while the lymphocyte ratio was significantly lower (Figure 3A). Red blood cell count, hemoglobin, and hematocrit were significantly lower in cluster 1 (p < 0.05 for all), and other erythrocyte-related parameters also showed significant differences between the clusters (Figure 3B). Platelet count tended to be lower in cluster 1 (Figure 3C), and immature granulocytes, including IG, promyelocytes, and myelocytes, were significantly higher in cluster 1 (Figure 3D, p < 0.05 for all). These results indicate that CBC data at diagnosis can stratify CML patients into groups with significantly different subsequent treatment responses.

**Figure 2.**
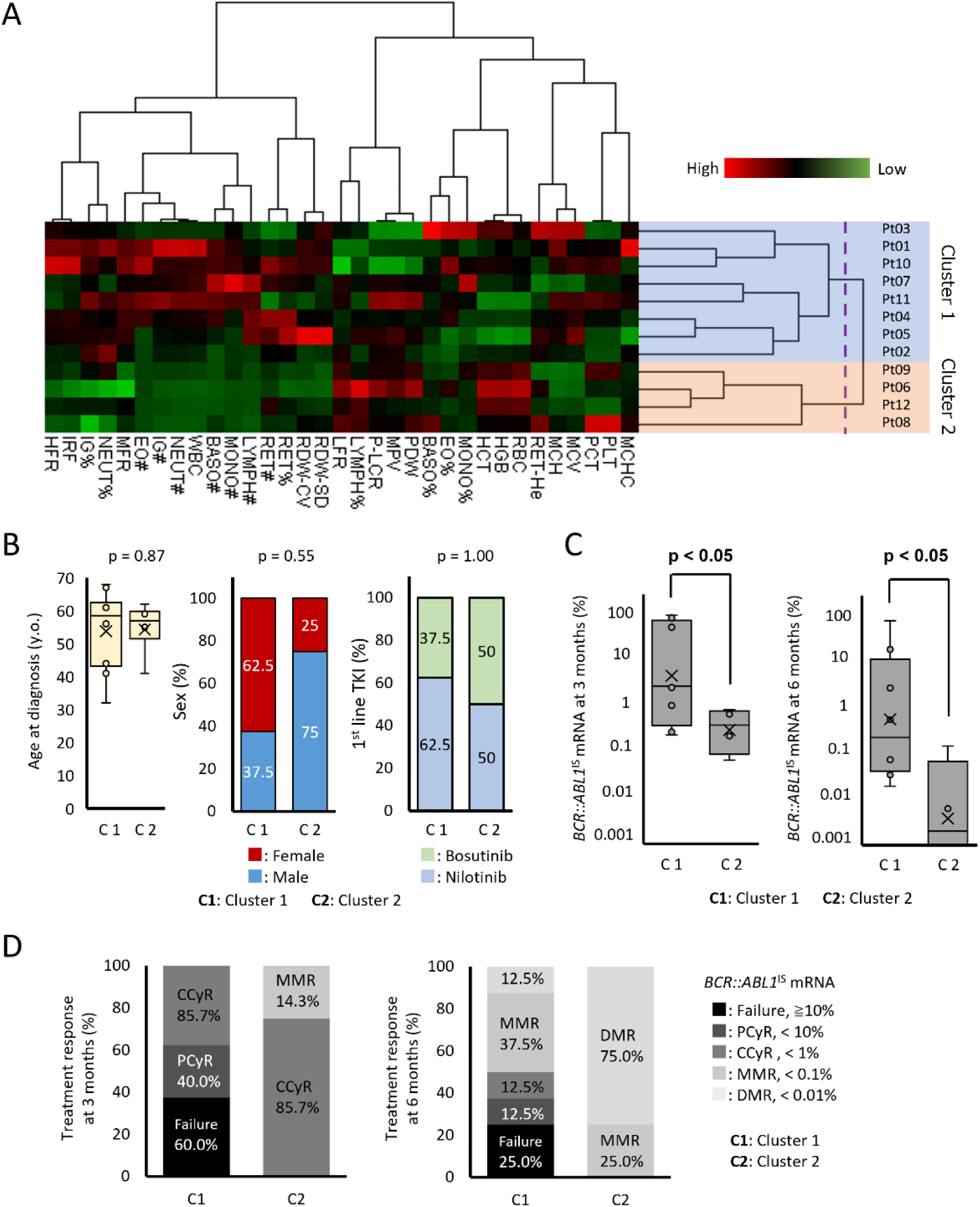
Stratification of patients with CML using CBC data. A. Hierarchical clustering analysis stratified CML patients into two groups: Cluster 1 and Cluster 2. B. Comparison of age, gender, and first-line TKI between the two clusters. C. Comparison of *BCR::ABL1*^IS^ mRNA levels at three- and six-months post-treatment initiation. D. Treatment responses between cluster 1 and 2. PCyR, Partial Cytogenetic Response; CCyR, Complete Cytogenetic Response; MMR, Major Molecular Response; DMR, Deep Molecular Response; Failure, those who did not achieve PCyR.

**Figure 3.**
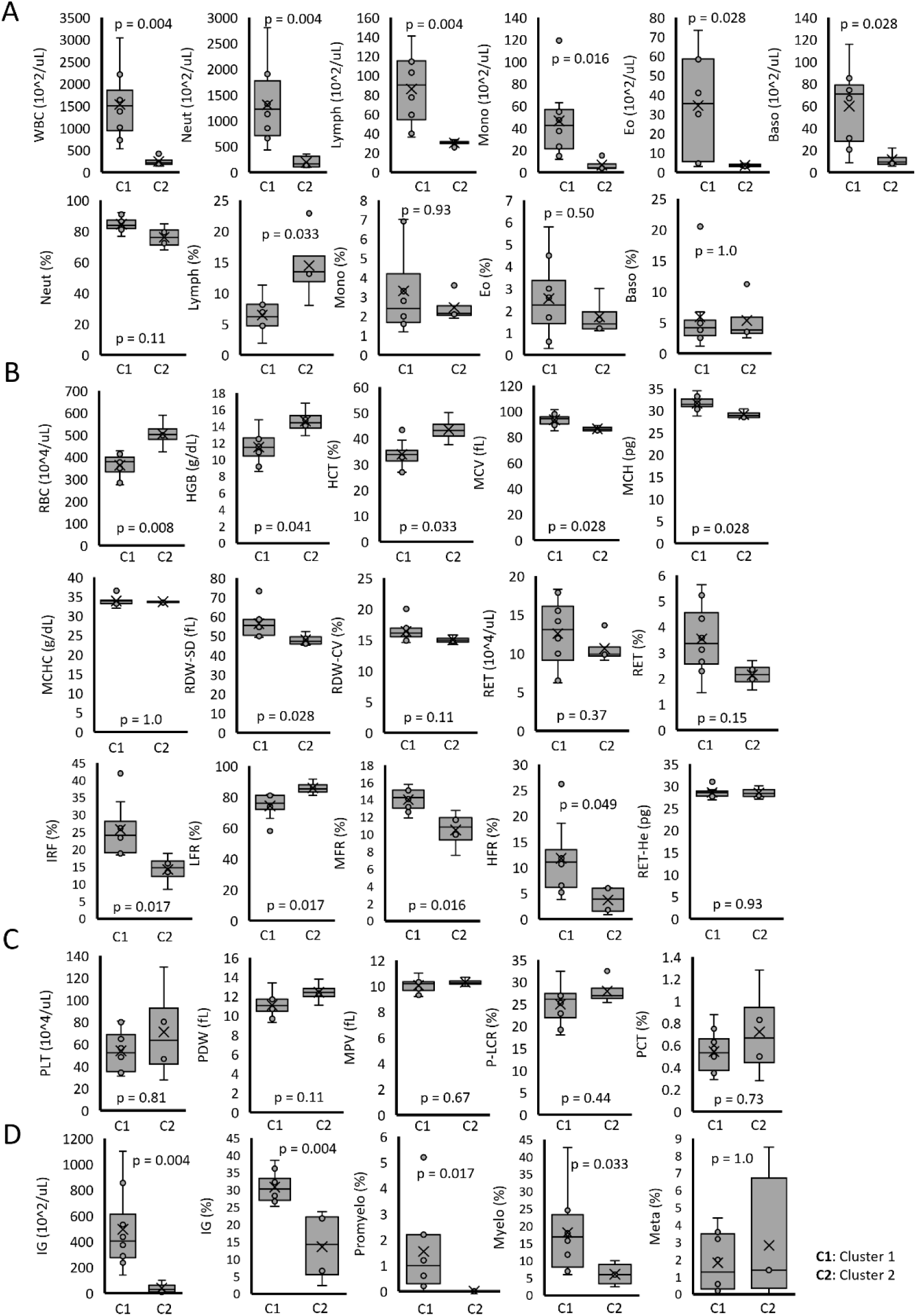
Comparison of CBC data between the two clusters. A. White blood cell-related parameters, B. red blood cell-related parameters, C. platelet-related parameters, and D. immature cell-related parameters were compared between cluster 1 and 2.

### Bone marrow progenitor cells differ between the patient groups with different treatment responses

To further investigate the underlying causes of the differences in CBC, we performed flow cytometric analysis of bone marrow mononuclear cells obtained at diagnosis. Among CD45⁺ cells, CD34^+^, CD38^-^, CD90^+^, and CD45RA^-^ cells were defined as HSCs; CD34⁺, CD38⁻, CD90⁻, and CD45RA⁻ cells as MPPs; and CD34⁺, CD10⁺ cells as CLPs. Furthermore, among CD45⁺, CD34⁺, CD38⁺, and CD10⁻ cells, those positive for CD135 and negative for CD45RA were defined as CMPs, those positive for both CD135 and CD45RA as GMPs, and those negative for both CD135 and CD45RA as MEPs (Figure 4A). When the proportions of each cell population relative to CD45⁺ cells were calculated, all HSPC populations tended to be increased in cluster 1 cases, with HSCs and GMPs being significantly higher in cluster 1 compared to cluster 2 (Figure 4B, p < 0.05 for both). Additionally, the ratio of CMP to CLP tended to be higher in cluster 1, and notably, the ratio of GMP to MEP was significantly higher in cluster 1 than in cluster 2 (Figure 4C, p < 0.01).

**Figure 4.**
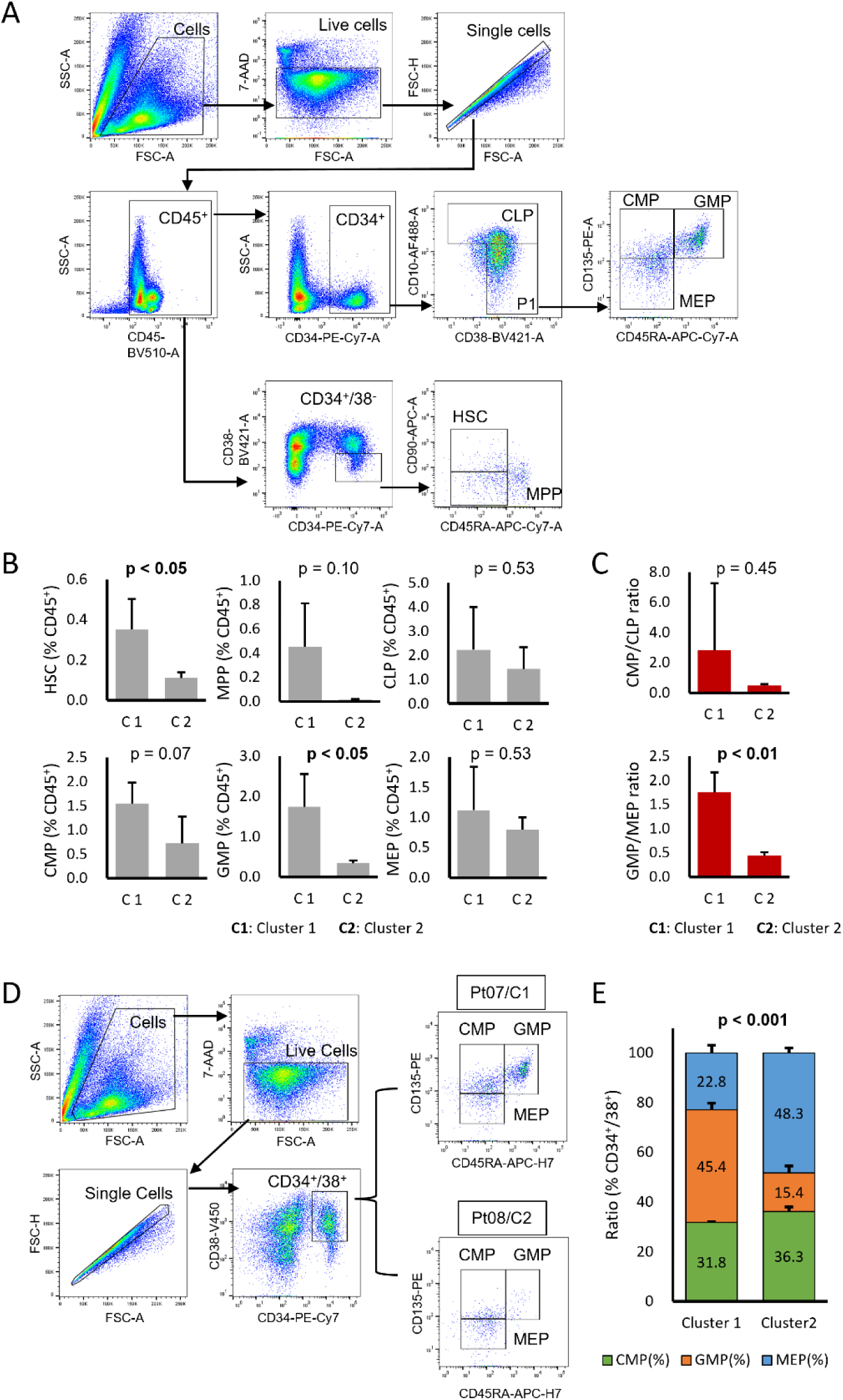
Flow cytometric analysis of bone marrow cells from patients with CML. A. The gating strategy defining HSPCs. B. Comparison of ratio of HSC, MPP, CLP, CMP, GMP, and MEP within CD45^+^ leukocytes between the two clusters. C. Comparison of ratio of CMP to CLP and GMP to MEP between the two clusters. D. The simple gating strategy defining CMP, MEP, and GMP. D. Comparison of CMP, MEP, and GMP within CD34^+^/38^+^ cells between the two clusters. HSC, hematopoietic stem cell; MPP, Multi Potent Progenitor; CLP, Common Lymphoid Progenitor; CMP, Common Myeloid Progenitor; GMP, Granulocyte-macrophage progenitor; MEP; Megakaryocyte-erythroid progenitor.

Considering that the GMP/MEP ratio may serve as a predictive marker for treatment response, we further analyzed these ratios using a simplified marker set. Among CD34⁺, CD38⁺ cells, CD135⁺, CD45RA⁻ cells were defined as CMPs, CD135⁺, CD45RA⁺ cells as GMPs, and CD135⁻, CD45RA⁻ cells as MEPs (Figure 4D). Comparison of these cell ratios between clusters revealed significant differences (p < 0.01), with no substantial difference in the proportion of CMPs, but a higher proportion of GMPs (45.4% vs 15.4%) and a lower proportion of MEPs (22.8% vs 48.3%) in cluster 1 compared to cluster 2 (Figure 4E).

These findings suggest that the differentiation balance of HSPCs differs between patient groups with distinct treatment responses.

### Artificial intelligence-driven flow cytometric analysis of peripheral leukocytes from patients with CML

We have previously reported that AI-driven ghost cytometry (GC)^21,22^, which captures subtle morphological information of cells in a label-free manner, enables discrimination of CML cells^18,23^. In this study, we investigated whether GC could comprehensively and conveniently detect alterations in CBC profiles arising from the differentiation balance of HSPCs, thereby enabling the prediction of treatment efficacy in CML patients using peripheral blood samples at diagnosis. First, peripheral blood leukocytes from CML patients at diagnosis and healthy donors were analyzed by GC, and discrimination scores (F1 score) based on AI analysis were calculated. Subsequently, the correlation between the F1 score at diagnosis and *BCR::ABL1*^IS^ mRNA levels at 3 and 6 months after treatment initiation was evaluated. Cells were classified into total leukocytes, lymphocytes, and granulocytes using side scatter (SSC)-forward scatter (FSC) scattergrams, and each fraction was analyzed separately (Figure 5A).

**Figure 5.**
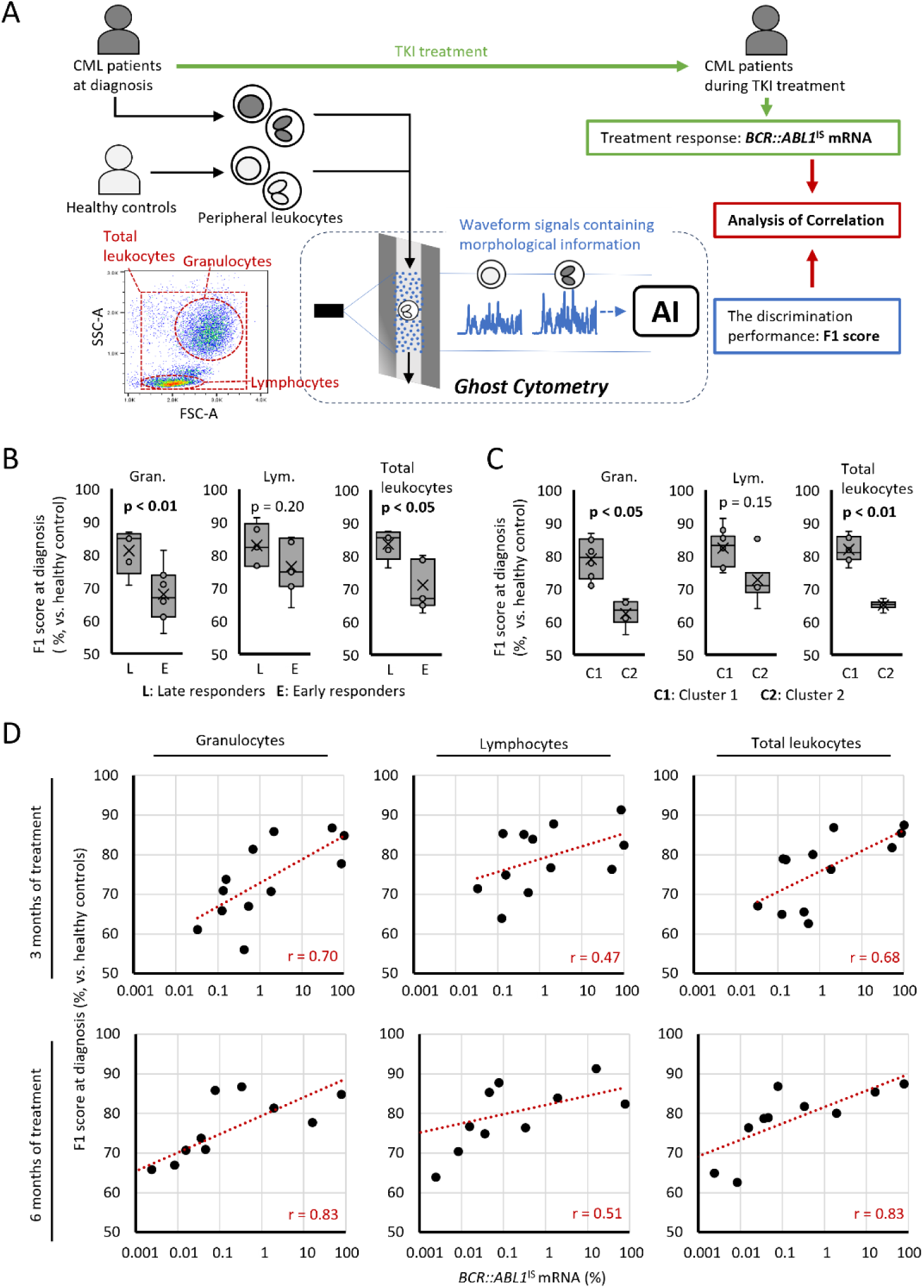
Ghost cytometric analysis of cells from patients with CML. A. Overview of the analysis. Peripheral blood leukocytes from CML patients at diagnosis and healthy controls were evaluated using GC, and their discrimination performance was calculated as the F1 score. The efficacy of TKI treatment in CML patients was assessed based on *BCR::ABL1*^IS^ mRNA, and the correlation with the F1 score was further analyzed. Gating of granulocytes, lymphocytes and total leukocytes using FSC-SSC scattergrams. Comparison of F1 scores between B. late and early responders and C. Cluster 1 and 2. Comparisons were made for granulocytes, lymphocytes, and total leukocytes fractions. D. The correlation between *BCR::ABL1*^IS^ mRNA at three and six months post-treatment initiation and F1 scores at diagnosis were assessed using Pearson’s rank correlation coefficient (r).

As a result, in both the early/late responder groups and cluster 1/2 groups defined in Figures 1A and 2A, the groups with poor treatment response (late responders and cluster 1) showed significantly higher F1 scores at diagnosis in the total leukocyte and granulocyte fractions (Figure 5B, C, p < 0.05 for all). Furthermore, the discrimination score at diagnosis showed a strong positive correlation (r = 0.68 – 0.83) with *BCR::ABL1*^IS^ mRNA levels at 3 and 6 months after treatment initiation, particularly in the total leukocyte and granulocyte fractions (Figure 5D).

These findings suggest that GC is a promising tool for the convenient analysis of peripheral blood samples and for predicting treatment response in CML.

## Discussion

This study suggests that the differentiation balance of bone marrow HSPCs in CML patients is associated with treatment sensitivity and resistance. Furthermore, changes in this differentiation balance alter the peripheral blood CBC profile, which may be useful for predicting treatment response and is consistent with our previous study^24^. Comprehensive detection of these changes using GC may provide a valuable tool for predicting treatment efficacy in CML (Figure 6).

**Figure 6.**
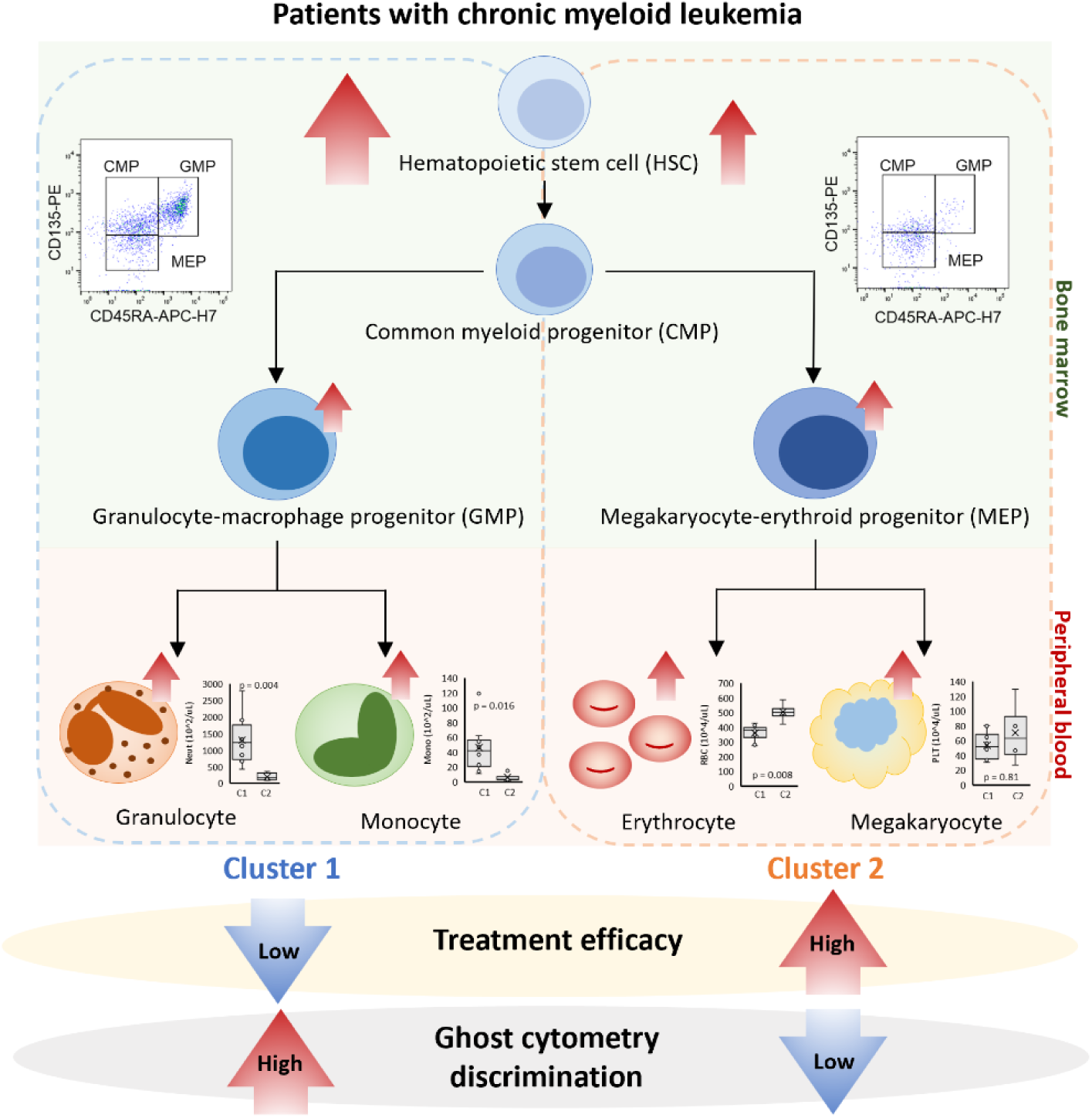
Schematic overview of the research.

Currently, multiple adenosine triphosphate-competitive TKIs^6^ and myristoyl pocket-targeting TKIs^25,26^ are available as therapeutic options for CML, and the ability to predict treatment efficacy at diagnosis could provide important information to guide therapeutic strategies. While myristoyl pocket-targeting asciminib is expected to become a useful choice due to its efficacy and tolerability in many CML patients^27,28^, considering the economic burden on patients and healthcare systems, those identified as highly treatment-sensitive in this study may benefit from the proactive selection of generic-available first- or second-generation TKIs. Selecting the optimal TKI for each patient based on such indicators may enable early achievement of deep remission and improve the rate of TFR. Moreover, previous study showed that treatment response at 6 months predicts DMR^29^, and higher DMR rate associate with successful TFR^30^. Our study demonstrated the ability to predict treatment response at 6 months, which suggests the potential to also forecast subsequent DMR and TFR.

In cases with strong *BCR::ABL1* activity and a large number of highly proliferative LSCs, it was considered that the number of cells in the HSC fraction increased, leading to an increase in peripheral blood leukocyte count. In our results, patients in cluster 1 with poor treatment response showed an increase in HSCs and associated progenitor cells. These groups also exhibited significantly elevated peripheral blood leukocyte counts, suggesting that leukocyte count at diagnosis should be considered when predicting early treatment response. However, leukocyte count can fluctuate due to physiological responses, infections, inflammatory diseases, and endocrine/metabolic disorders, and delayed diagnosis of CML may also contribute to leukocytosis. Therefore, while leukocyte count at diagnosis may be a useful indicator for predicting treatment response, it is difficult to rely on it as a single parameter.

Clustering analysis of CBC revealed differences in peripheral blood profiles between patient groups with distinct treatment responses. This was considered to be due to differences in the balance of differentiation from CMP to GMP or MEP. Differentiation from CMP to GMP is strongly regulated by *Purine-rich Box 1* (*PU.1*)^31,32^, while differentiation from CMP to MEP is regulated by *GATA Binding Protein 1* (*GATA-1*)^33^. CMPs are in a lineage priming state with low expression of *PU.1* and *GATA-1*; when *PU.1* predominates, it suppresses *GATA-1* and promotes differentiation toward GMP, whereas when *GATA-1* predominates, it suppresses *PU.1* and promotes differentiation toward MEP^16,34,35^. In CML cases, this balance of CMP differentiation may be a factor determining treatment sensitivity and resistance, but further studies are needed to validate this mechanism.

GC is a technology that enables label-free detection of subtle cellular structural changes and AI-based cell discrimination without the need for antibody staining or other cell processing^21,22^. We have previously reported that GC can distinguish CML cells from normal cells by capturing mitochondrial morphological changes^23^. Furthermore, by training AI with GC data from immature granulocytes with low CD16 expression^36^, the detection accuracy for CML cells was improved, and the immaturity of CML granulocytes was identified as a key factor for AI-based discrimination^18^. In this study, the patient groups with poor treatment response showed increased numbers of immature leukocytes such as IGs, promyelocytes, and myelocytes, which likely contributed to higher GC discrimination scores. Additionally, the increase in immature leukocytes may be due to enhanced differentiation from CMP to GMP, resulting in increased immature granulocytes. The greater difference in discrimination scores in the granulocyte fraction compared to the lymphocyte fraction (Figure 5B, C) is consistent with our interpretation. Recently, neutrophil abundance and granulocytic maturation have been reported to be associated with sustained TFR^37^. Since our prediction of treatment response is influenced by granulocytic maturation, it may also have the potential to predict not only early treatment response but subsequent TFR.

This study has several limitations. First, the number of cases analyzed was small, and further validation with larger cohorts is required to confirm these findings. For the same reason, the statistical power is limited, and some results may be under- or overestimated. In this study cohort, all patients received second or third-generation TKIs, or asciminib. Therefore, the potential bias in treatment response due to the type of TKI is considered minimal. However, further validation in larger cohorts under consistent treatment conditions is required. Moreover, the follow-up period in this study was limited to 6 months, and future studies are needed to assess treatment response at 12 and 24 months and its association with TFR.

In conclusion, this study suggests that differences in the differentiation balance of bone marrow HSPCs exist between CML patients with distinct treatment responses, leading to changes in peripheral blood profiles. Multivariate analysis of peripheral blood cells and AI-driven GC may enable convenient and accurate prediction of treatment efficacy at diagnosis, providing valuable information for the development of CML treatment strategies.

## Authors’ Contributions

K.S. designed, performed, and analyzed experiments and contributed to writing the manuscript. N.W., Y.T., T.I., and S.K. collected clinical specimens and information and contributed to clinical and scientific discussions. K.Y. developed a prototype of ghost cytometry and provided the methodology. M.A. supervised the research and contributed to clinical and scientific discussions. T.T. designed and supervised the research, collected clinical specimens and information, and contributed to writing the manuscript. All authors approved the final manuscript.

## Acknowledgement

We thank the Laboratory of Cell Biology and Molecular and Biochemical Research in Biomedical Research Core Facilities, Juntendo University Graduate School of Medicine, for their technical assistance. We also thank the researchers and engineers at Sysmex Corporation for their contributions to the development of a prototype of ghost cytometry and for providing the associated methodology.

## Data Availability

The datasets generated and/or analyzed during the current study are available from the corresponding author upon reasonable request.

